# The Metabolic Triad: Trends and Determinants of the Isolated and Combined Presence of Hypertension, Obesity, and Diabetes in Peru

**DOI:** 10.1101/2024.04.22.24306195

**Authors:** Víctor Juan Vera-Ponce, Fiorella E. Zuzunaga-Montoya, Luisa Erika Milagros Vásquez-Romero, Joan A. Loayza-Castro, Cori Raquel Iturregui Paucar, Enrique Vigil-Ventura

**Author notes:** Correspondence Víctor Juan Vera-Ponce. Luisa Erika Milagros Vásquez Romero.

## Abstract

**Introduction:** **In** recent years, the presence of non-communicable diseases, particularly hypertension (HTN), obesity, and type 2 diabetes mellitus (T2DM), has emerged as a significant threat to global public health.

**Objective:** To determine the trends and factors associated with these diseases’ isolated and combined presence in both dual and triple forms (metabolic triad).

**Methods:** Secondary data analysis from the Demographic and Family Health Survey for 2014–2022. The primary variable had seven categories, ranging from isolated states to triple combinations. Given the polytomous nature of the dependent variable, multinomial logistic regression models were constructed. The results of these models are presented in the form of adjusted odds ratios, accompanied by their 95% confidence intervals (95% CI).

**Results:** Among the isolated forms, obesity was the most prevalent comorbidity (15.35%), while T2DM was the least common (1.14%). Among the combined forms, the most frequent was obesity and HTN (6.40%). The prevalence of the metabolic triad was low (0.84%). The factors commonly related to each were female sex, age, urban residency, region, economic level, daily smoking, alcohol consumption, and altitude.

**Conclusions:** An increasing prevalence of HTN, T2DM, and obesity was found, both individually and in their various combinations. Furthermore, the complexity and sometimes contradictory nature of the associated factors, which can increase or decrease the presence of these metabolic diseases, reflect the need for a personalized approach to prevention and treatment.

## Introduction

In recent years, the presence of non-communicable diseases, especially arterial hypertension (HTN), obesity, and type 2 diabetes mellitus (T2DM), in worldwide population well-being has become increasingly difficult to ignore ^(1)^. While these conditions alone undermine an individual’s quality of life, their chronic nature likewise increases susceptibilities to long-term severe complications involving cardiovascular and renal dysfunctions as well as earlier mortality than average through multiple pathways, as corroborated by various scientific reports ^(2,3)^.

The increasing prevalence of these diseases is not uniformly distributed across different regions and populations, showing marked geographical and demographic disparities. Recent studies have found that although high-income nations have seen stabilization in prevalence due to strengthened public health policies and treatment access, low- and middle-income countries continue experiencing rising prevalence attributable to ongoing urbanization, lifestyle and dietary shifts, and an aging population ^(4–6)^.

These diseases have been noted to involve shared risk factors like physical inactivity, unhealthy diets, and genetic predisposition, which regularly result in their concurrent presentation—further compounding their effect on both individual and public health ^(7–9)^. Recent research has conclusively shown that concurrently facing HTN, obesity, and T2DM can drastically elevate one’s risks of infirmity and death far beyond what any such condition could singularly, emphasizing the need for preventative and therapeutic efforts aimed at curbing the progression of such metabolic dysfunctions ^(10–14)^.

In Peru, although each disease has been investigated in isolation ^(15,16)^, evidence of their combinations is still scarce. To craft more targeted and responsive public health strategies, this study seeks to gain nuanced insight into the trends and interconnected factors underlying the regional prevalence of these illnesses, given the importance of tailoring approaches to the distinct circumstances of local communities.

## Methods

### Design

An analytical cross-sectional study was conducted through the secondary analysis of information obtained from the Demographic and Family Health Survey (ENDES) provided by the National Institute of Statistics and Informatics (INEI) for the period 2014 to 2022 ^(17)^. This analysis was carried out by the STROBE (Strengthening the Reporting of Observational Studies in Epidemiology) guidelines, aiming to ensure high quality and transparency in the reportability of observational studies ^(18)^.

### Population, Eligibility Criteria, and Sample

The original study focused on Peruvian individuals aged 15 to 99 years, covering urban and rural areas across the country’s 24 departments. ENDES’s sampling methodology was based on a probabilistic, stratified, two-stage design differentiating urban and rural areas.

For this study, only individuals aged 20 years and older were included, following the standard definitions of adult subjects only. Individuals without measured blood pressure, weight, or height were excluded. Additionally, limits were established based on thresholds used in previous studies to ensure the inclusion of valid blood pressure measurements for analysis. Specifically, Systolic Blood Pressure (SBP) measurements in the 70 mmHg to 270 mmHg and Diastolic Blood Pressure (DBP) from 50 mmHg to 150 mmHg were considered acceptable. Measurements not meeting these validity criteria were excluded from the study ^(19)^.

### Variables and Measurement

The main variable had seven categories:

□ Isolated HTN: presence of SBP ≥ 140 mmHg or DBP ≥ 90 mmHg and by self-report.
□ Isolated T2DM: the presence of diabetes by self-report only.
□ Isolated Obesity: presence of a Body Mass Index (BMI) of ≥ 30 Kg/m2 only.
□ Combination of HTN and T2DM (HD): if HTN and T2DM were present together.
□ Combination of HTN and obesity (HO): if HTN and Obesity were present.
□ Combination of T2DM and obesity (DO): if T2DM and Obesity were present together.
□ Combination of all three diseases (metabolic triad): if HTN, T2DM, and obesity were present.

The study analyzed participants’ characteristics, including sex (female or male), age (categorized into three groups: 20-44 years, 45-59 years, and 60 years or older), residence region (Metropolitan Lima, Rest of the coast, Highlands or Jungle), educational level (none/primary or secondary/higher), wealth index (very poor/poor, middle or rich/very rich), residence area (urban or rural), habits (daily smoker, alcohol consumption in the last 30 days), and residence altitude (categorized into four ranges: 0 to 499 meters above sea level, 500 to 1499 meters, 1500 to 2999 meters, or 3000 meters or more).

### Procedures

To determine the Body Mass Index, one takes the participant’s weight in kilograms and divides it by the square of their height in meters, thereby calculating this metric. Weight measurement was carried out using an electronic scale of 0.1 kg, ensuring participants were in light underwear and barefoot. In strict adherence to international and domestic protocols, the researchers precisely gauged height via a wooden stadiometer, calibrating measurements to the nearest millimeter.

For blood pressure measurement, participants sat calmly, with the right arm aligned at heart level on a horizontal support. An initial measurement was taken to stabilize the subject’s cardiovascular conditions after resting for five minutes, followed two minutes later by a second assessment. The mean value of these two readings, both for SBP and DBP, was calculated and used in subsequent analyses. This approach aims to minimize transient variations in blood pressure, providing a more accurate and representative estimate. The measurement was carried out following a uniform protocol, using an OMRON digital sphygmomanometer, model HEM-713, and two different cuff sizes were used to accommodate variations in participants’ arm size, one for standard-sized arms (220–320 mm) and another for larger arms (330–430 mm).

### Statistical Analysis

The present study utilizes the R program for conducting various statistical analyses. It has been considered throughout the process that it involves complex sampling analysis, which implies special data treatment to obtain valid results.

Thus, the analysis was divided into three stages. First, a descriptive analysis of all variables involved in the study was carried out. In the second stage, a trend analysis was performed to observe the evolution of each comorbidity between 2014 and 2022. This analysis is conducted for each comorbidity individually and for the set of comorbidities, thus identifying whether there is an increase or decrease in the prevalence of comorbidities over time.

Finally, given the polychotomous nature of the dependent variable, multinomial logistic regression models were constructed. These models allow determining the factors independently associated with each evaluated comorbidity, compared to those not presenting any of these pathologies. The results of these models are presented as adjusted odds ratios (aOR) with their 95% confidence intervals (CI 95%).

### Ethical Considerations

This study is based on public data, available at no cost and without personal information that could identify participants ^(20)^. Thus, privacy is protected, and ethical risks are eliminated. For these reasons, the evaluation of an ethics committee was not required for this study.

## Results

We worked with a total of 309,652 participants. Descriptive characteristics are displayed in Table 1. In general,

**Table 1.** Demographic characteristics of participants.

| **Characteristic** | **n = 309,652** |
| --- | --- |
| **Sex** |  |
| Male | 149,712 (48.35%) |
| Female | 159,940 (51.65%) |
| **Group age** |  |
| 20 - 44 years | 171,190 (59.58%) |
| 45 - 59 years | 64,902 (22.59%) |
| 60 to more | 51,229 (17.83%) |
| **Educational Level** |  |
| No Level/Primary | 117,236 (37.86%) |
| Secondary/Superior | 192,416 (62.14%) |
| **Natural region** |  |
| Metropolitan Lima | 104,935 (33.89%) |
| Resy of coast | 79,280 (25.60%) |
| Mountain Range | 84,329 (27.23%) |
| Jungle | 41,107 (13.28%) |
| **Wealth index** |  |
| The poorest/Poor | 127,060 (41.03%) |
| Medium | 62,819 (20.29%) |
| Rich/Richest | 119,773 (38.68%) |
| **Area of residence** |  |
| Rural | 72,222 (23.32%) |
| Urbano | 237,429 (76.68%) |
| **Smoker Status** |  |
| No | 304,856 (98.45%) |
| Yes | 4,796 (1.55%) |
| **Alcohol consumption** |  |
| No | 104,046 (50.40%) |
| Yes | 102,396 (49.60%) |
| **Altitude** |  |
| 0 to 1499 | 220,179 (71.12%) |
| 1500 to 2499 | 20,976 (6.78%) |
| 2500 to 3499 | 43,163 (13.94%) |
| 3500 or more | 25,264 (8.16%) |
| **Metabolic diseases** |  |
| No | 187,821 (62.52%) |
| Obesity Only | 46,146 (15.36%) |
| HTN Only | 35,945 (11.97%) |
| T2DM Only | 3,418 (1.14%) |
| Obesity and T2DM | 1,730 (0.58%) |
| Obesity and HTN | 19,217 (6.40%) |
| T2DM and HTN | 3,607 (1.20%) |
| Obesity, T2DM, and HTN | 2,526 (0.84%) |
| n (%) |  |

Among isolated conditions, obesity was the most prevalent comorbidity (15.35%), while T2DM was the least common (1.14%). Among combined conditions, the most frequent was the combination of obesity and HTN (6.40%). The prevalence of the metabolic triad was low (0.84%).

Trend analysis showed fluctuations over time in the prevalence of isolated forms of obesity and T2DM, with a decrease in HTN. However, there was an increase in OD, OH, and DH prevalence in recent years, along with a more significant rise in the metabolic triad.

The multivariable analysis presented in Table 2 identified factors distinctly associated with each condition and their combinations. In the case of isolated obesity, associated factors included being female, older age, higher educational level, residing in coastal and Andean regions outside the capital, belonging to a middle economic level, living in urban areas, being a daily smoker, alcohol consumption, and greater altitude, the latter acting as a protective factor. On the other hand, HTN showed associations with age, living in specific regions that appear to exert a protective effect, having a high economic level, residing in urban areas, being a daily smoker, alcohol consumption, and altitude. For T2DM, associated factors were age, educational level, living in specific regions that offer a protective effect, residential area, alcohol consumption (also with a protective effect), and altitude.

**Table 2.** Multivariable regression analysis of the factors associated with HTN, T2DM, Obesity, and their combinations.

| Characteristic | Only Obesity |  | Only HTN |  | Only T2DM |  | Obesity + T2DM |  | Obesity + HTN |  | T2DM + HTN |  | Obesity + HTN + T2DM |  |
| --- | --- | --- | --- | --- | --- | --- | --- | --- | --- | --- | --- | --- | --- | --- |
|  | aOR | 95% CI | aOR | 95% CI | aOR | 95% CI | aOR | 95% CI | aOR | 95% CI | aOR | 95% CI | aOR | 95% CI |
| **Sex** |  |  |  |  |  |  |  |  |  |  |  |  |  |  |
| Male | — | — | — | — | — | — | — | — | — | — | — | — | — | — |
| Female | **1.79** | **1.74, 1.84** | **0.69** | **0.66, 0.71** | 0.99 | 0.91, 1.07 | **1.65** | **1.48, 1.84** | **0.92** | **0.88, 0.96** | **0.82** | **0.75, 0.90** | **1.19** | **1.09, 1.30** |
| **Group age** |  |  |  |  |  |  |  |  |  |  |  |  |  |  |
| 20 – 44 years | — | — | — | — | — | — | — | — | — | — | — | — | — | — |
| 45 – 59 years | **1.66** | **1.61, 1.71** | **3.78** | **3.65, 3.93** | **3.68** | **3.37, 4.02** | **4.85** | **4.33, 5.44** | **4.63** | **4.43, 4.84** | **4.17** | **3.76, 4.62** | **5.61** | **5.04, 6.25** |
| 60 to more | **1.27** | **1.22, 1.34** | **9.87** | **9.47, 10.3** | **6.55** | **5.93, 7.24** | **6.40** | **5.57, 7.34** | **6.66** | **6.31, 7.02** | **11.3** | **10.2, 12.6** | **14.4** | **12.9, 16.0** |
| **Educational Level** |  |  |  |  |  |  |  |  |  |  |  |  |  |  |
| No Level/Primary | — | — | — | — | — | — | — | — | — | — | — | — | — | — |
| Secondary/Superior | **1.12** | **1.08, 1.15** | 1.03 | 0.99, 1.07 | **1.13** | **1.03, 1.25** | **1.16** | **1.03, 1.32** | **1.06** | **1.01, 1.11** | **0.85** | **0.77, 0.94** | **1.24** | **1.11, 1.37** |
| **Natural region** |  |  |  |  |  |  |  |  |  |  |  |  |  |  |
| Metropolitan Lima | — | — | — | — | — | — | — | — | — | — | — | — | — | — |
| Resy of coast | **1.22** | **1.18, 1.26** | **0.74** | **0.71, 0.77** | **0.8** | **0.73, 0.89** | **1.24** | **1.09, 1.41** | **1.09** | **1.04, 1.14** | 0.93 | 0.84, 1.03 | 1.00 | 0.90, 1.11 |
| Mountain Range | **1.37** | **1.21, 1.56** | **0.32** | **0.29, 0.37** | **0.33** | **0.24, 0.47** | **4.85** | **2.83, 8.33** | **0.28** | **0.24, 0.33** | **1.95** | **1.22, 3.10** | **0.21** | **0.14, 0.32** |
| Jungle | 0.98 | 0.93, 1.02 | **0.64** | **0.60, 0.68** | **0.64** | **0.55, 0.74** | **1.83** | **1.53, 2.18** | **0.54** | **0.50, 0.59** | 1.08 | 0.93, 1.27 | **0.83** | **0.70, 0.99** |
| **Wealth index** |  |  |  |  |  |  |  |  |  |  |  |  |  |  |
| The poorest/Poor | — | — | — | — | — | — | — | — | — | — | — | — | — | — |
| Medium | **1.14** | **1.10, 1.18** | 0.99 | 0.94, 1.04 | 0.89 | 0.79, 1.01 | **1.56** | **1.30, 1.87** | **1.32** | **1.24, 1.40** | **1.39** | **1.20, 1.63** | **1.18** | **1.01, 1.37** |
| Rich/Richest | 1.02 | 0.98, 1.05 | **0.81** | **0.77, 0.85** | 0.92 | 0.83, 1.03 | **2.01** | **1.70, 2.38** | **1.07** | **1.01, 1.14** | **1.95** | **1.70, 2.25** | **1.37** | **1.20, 1.58** |
| **Area of residence** |  |  |  |  |  |  |  |  |  |  |  |  |  |  |
| Rural | — | — | — | — | — | — | — | — | — | — | — | — | — | — |
| Urbano | **1.93** | **1.84, 2.02** | **1.29** | **1.22, 1.36** | **2.09** | **1.80, 2.43** | **1.59** | **1.30, 1.95** | **1.9** | **1.76, 2.05** | **1.76** | **1.48, 2.08** | **3.06** | **2.51, 3.74** |
| **Smoker Daily** |  |  |  |  |  |  |  |  |  |  |  |  |  |  |
| No | — | — | — | — | — | — | — | — | — | — | — | — | — | — |
| Yes | **1.36** | **1.25, 1.47** | **1.26** | **1.15, 1.39** | 1.15 | 0.90, 1.46 | 0.80 | 0.55, 1.17 | **1.28** | **1.14, 1.43** | 1.03 | 0.79, 1.34 | **0.64** | **0.45, 0.90** |
| **Alcohol consumption** |  |  |  |  |  |  |  |  |  |  |  |  |  |  |
| No | — | — | — | — | — | — | — | — | — | — | — | — | — | — |
| Yes | **1.11** | **1.08, 1.14** | **1.04** | **1.01, 1.07** | **0.87** | **0.81, 0.94** | **1.53** | **1.37, 1.69** | **1.22** | **1.17, 1.26** | 0.92 | 0.85, 1.00 | 0.95 | 0.87, 1.03 |

| Altitude |  |  |  |  |  |  |  |  |  |  |  |  |  |  |
| --- | --- | --- | --- | --- | --- | --- | --- | --- | --- | --- | --- | --- | --- | --- |
| 0 – 1499 | — | — | — | — | — | — | — | — | — | — | — | — | — | — |
| 1500 – 2499 | 0.55 | 0.49, 0.62 | 2.13 | 1.92, 2.37 | 1.78 | 1.34, 2.37 | 0.19 | 0.11, 0.33 | 2.33 | 2.03, 2.67 | 0.39 | 0.25, 0.61 | 2.05 | 1.48, 2.84 |
| 2500 – 3499 | 0.45 | 0.39, 0.51 | 2.24 | 1.97, 2.55 | 1.37 | 0.95, 1.97 | 0.20 | 0.12, 0.35 | 1.86 | 1.56, 2.21 | 0.30 | 0.19, 0.48 | 3.88 | 2.53, 5.94 |
| 3500 or more | 0.45 | 0.39, 0.52 | 2.55 | 2.23, 2.92 | 2.26 | 1.55, 3.28 | 0.25 | 0.14, 0.44 | 1.60 | 1.32, 1.94 | 0.42 | 0.26, 0.69 | 2.74 | 1.70, 4.41 |

\*each factor has been independently adjusted for sex, group age, educational level, region, area of residence, wealth index, daily smoking, alcohol consumption, consumption of fruits and vegetables, and altitude
aRP: Adjusted Prevalence ratio. 95% CI: 95% confidence interval
Source: self-made

OD shared similar associations, including being female, age, educational level, region of residence, economic level, urban residency, alcohol consumption, and altitude, with the latter acting as a protective factor. For the OH combination, associated factors were being female (protective effect), age, educational level, residing in regions with a protective effect, residential area, being a daily smoker, alcohol consumption, and altitude. In the case of OD, factors included being female (protective effect), age, having a high educational level (protective), living in the mountain region, belonging to a middle or high economic level, residing in urban areas, and altitude (protective effect).

Finally, for the metabolic triad, related factors encompassing being female, age, higher educational level, living in different natural regions, economic level, urban residency, and altitude.

## Discussion

### Prevalence and Trends of Obesity, HTN, T2DM in Isolation and Their Combinations

The increasing simultaneous coexistence of HTN, T2DM, and obesity reported elsewhere globally can likely be ascribed to an assortment of interconnected socioeconomic determinants ^(3,11–14,21)^. Firstly, while population aging and shifts towards sedentary lifestyles and diets high in fats and refined carbohydrates associated with increased urbanization could partially explain, studies have also linked changes in underlying risk factors such as growing obesity and metabolic disorders among specific demographics to rising disease prevalence in numerous nations The interaction of certain lifestyle habits can increase susceptibility to metabolic health issues ^(22,23)^.

Secondly, the joint existence of these conditions could represent an oblique manifestation of metabolic syndrome, which constitutes a grouping of cardiovascular risk determinants such as insulin resistance and endothelial impairment. While the growing frequency of these disease combinations could signify an emerging rise in the frequency of metabolic syndrome itself, research from the past few years shows higher rates of co-occurrence for the disorders involved ^(24,25)^.

Thirdly, improvements in detecting and diagnosing these diseases over time could contribute to the increasing rates of combined diseases. Greater awareness and better screening practices in healthcare might be identifying more individuals living with multiple chronic conditions ^(26,27)^.

Fourthly, pharmacological interactions and treatment complications may play a role in the rise of disease combinations. For instance, certain medications used for treating HTN may influence body weight or glucose control, thus contributing to obesity or the development of T2DM ^(28)^.

Fifthly, inequitable access to crucial socioeconomic pillars, including healthcare, education, and health management resources, is thought to potentially impact the uneven distribution of concurrent diseases across different segments of society. Underprivileged populations are often most affected by chronic diseases and their combinations due to a lack of access to healthy foods, exercise spaces, and preventive healthcare ^(29,30)^.

Ultimately, the interplay between genetic and epigenetic elements could underlie why some groups may exhibit these sicknesses jointly or concurrently, perhaps illuminating why certain persons evolve numerous metabolic disturbances. In contrast, others appear exempt ^(31)^.

The increase in combinations of metabolic diseases, especially the triad of HTN, T2DM, and obesity, is a call to action for public health and medical research. This phenomenon makes evident the necessity for a holistic methodology within healthcare that addresses each disease independently and considers the interrelations between them and their common causal elements. This study underscores the need for proactive and inclusive public health strategies that cultivate healthy habits and provide fair access to medical care to curb the onset and worsening of interrelated chronic conditions.

### Associated Factors of Obesity, HTN, T2DM, and Their Combinations

Regarding the associated factors found, the complex and sometimes contradictory nature of factors that increase or decrease the presence of obesity, HTN, and T2DM, as well as their combinations, is highlighted.

One of the most intriguing findings is the role of the female sex, which is associated with a higher risk of isolated obesity but acts as a protective factor in OH and OD combinations. This suggests that the impact of sex on the risk of chronic diseases is multifaceted and may be mediated by biological, behavioral, and socioeconomic differences between men and women. These differences may be influenced by hormonal variations, differences in body composition and fat distribution, and social and behavioral factors, including disparities in access to healthcare, differences in dietary behaviors and physical activity, and the impact of cultural and gender norms ^(32,33)^.

Age is confirmed as a consistent risk factor across all conditions examined, reflecting the accumulation of adverse exposures over a lifetime and a natural decline in physiological resilience. This finding underscores the importance of early and life-stage-adapted preventive interventions that can mitigate the cumulative impact of risk factors and promote healthy aging ^(8,34–36)^.

The influence of geographic context on the risk of chronic diseases and their combinations points to the relevance of local environmental and socioeconomic factors. Regional variability in the risk of these conditions underscores the need for personalized public health approaches, considering each region’s specific characteristics to develop more effective intervention strategies ^(16,37,38)^.

Educational level and socioeconomic status emerge as critical determinants, with effects varying according to the specific health condition. This indicates that education and income affect health knowledge awareness and access to resources that can mitigate or exacerbate disease risk.

Educational level and socioeconomic status play a critical role in affecting health knowledge awareness and access to healthy resources. Education and income influence lifestyle choices, including diet, physical activity, and health services, involving the risk of developing chronic conditions ^(39–41)^.

Finally, alcohol consumption, smoking, and height show complex and variable associations with cardiovascular and metabolic health. These factors can significantly interact with other health determinants, suggesting that their effects on disease risk are not uniform and may be influenced by concurrent risk factors. This nuanced understanding highlights the need to consider the interdependence of health behaviors and other risk factors in developing chronic diseases and the importance of intervention strategies that address these complex interactions ^(42–45)^.

It is essential to add that the protective role of daily smoking for the presence of metabolic triad was surprising. This may not be plausible, as various studies have found the harmful role that smoking plays in the development of multiple diseases, including those studied in this research ^(46,47)^. Therefore, the explanation behind this could lie in the study design. Expressly, cross-sectional studies, by offering a snapshot of the population at a specific time, are limited in their ability to establish temporal sequences between exposure and outcome. Thus, while findings may suggest a relationship between risk or protective factors and specific health conditions, they cannot definitively confirm that one variable causes the other. In the case of smoking and its association with a reduced risk of metabolic triad, it is plausible that this result does not reflect a natural protective effect of tobacco. The explanation for this occurrence could be the presence of reverse causality bias, where health conditions influence the behaviors or characteristics observed, further complicating the interpretation of these findings ^(48)^. In this case, individuals with a prior diagnosis of one or more of these chronic conditions may have modified their lifestyle habits, including smoking, in response to their health status or may have been diagnosed at early stages due to increased medical surveillance. Thus, this specific result should be interpreted with caution, considering both the valuable perspective they offer on the interactions between multiple risk factors and health conditions and the inherent limitations of their cross-sectional design. Including these nuances in our discussion enriches the understanding of the findings and guides future research directions and the application of public health interventions.

### Contribution to the Field

By examining patterns in the prevalence of HTN, T2DM, and obesity separately and concurrently, this research furthers comprehension of how these conditions evolve individually and jointly over time. Long-term surveillance of these trends is crucial for planning and evaluating public health policies.

The study delves deeply into how an intricate web of interconnected risk factors contributes to the multifaceted nature of having multiple chronic diseases simultaneously by closely examining the intricate interplay between various long-term health conditions. This knowledge is vital for designing more effective preventive interventions. By highlighting the necessity for multidimensional preventative and regulatory tactics focusing on dietary habits, physical activity, and obesity administration, the investigation underscores how addressing multiple influences can dramatically cut the occurrence of HTN, T2DM, and their interconnected accompanying afflictions.

By providing updated data on the prevalence and trends of these chronic diseases, the study informs resource allocation within the health system for clinical care and prevention, particularly in the most affected geographical areas and populations. The study’s results underscore the need for comprehensive public health initiatives that holistically tackle interconnected chronic illnesses rather than focusing on individual conditions in a silo, which is pivotal for enhancing a population’s aggregate health status.

Finally, publishing this data can raise awareness about the magnitude and seriousness of metabolic diseases and the importance of lifestyle factors in their development, encouraging the population to adopt healthy behaviors.

### Study Limitations

First, as previously mentioned, given the study’s cross-sectional nature, it only provides data at a specific time. Therefore, it cannot establish causality or temporal sequences between exposure and outcome. This limits the study’s ability to identify causal relationships between risk factors and diseases. Second, the analysis is based on secondary data collected by ENDES, which may introduce limitations in terms of information accuracy and data collection methodology not explicitly designed for this study.

Third, measurements of blood pressure, height, and weight were taken on a single occasion, which may not accurately reflect the chronic state or long-term control of these conditions, as guidelines even recommend doing so on up to two occasions to ensure results. Fourth, there may be unmeasured or inadequately adjusted confounding factors in the multivariable analysis that could influence the associations between risk factors and diseases. Finally, although ENDES is a national survey, subgroups of the population may be underrepresented or not represented in the sample.

## Conclusions

An increasing prevalence of HTN, T2DM, and obesity was found, both individually and in their various combinations. Moreover, the complexity and sometimes contradictory nature of the associated factors that can increase or decrease the presence of these metabolic diseases are shown, reflecting the need for a personalized approach to prevention and treatment. This complexity underscores the importance of a comprehensive public health approach that considers the interactions between multiple chronic conditions and the shared and unique risk factors for each disease.

Future research must be directed towards longitudinal studies to overcome the cross-sectional design limitations and explore causal relationships more deeply. These studies should strive to understand better how risk factors interact with each other and how effective interventions addressing the coexistence of multiple chronic conditions can be designed. Additionally, strengthening public health interventions that address the social and behavioral determinants of health, with particular attention to personalization based on identified risk factors, is recommended. Promoting healthy lifestyles, equitable access to preventive medical care, and appropriate health education are fundamental to mitigating the rising prevalence of these chronic conditions.

## Acknowledgments

A special thanks to the members of the Tropical Diseases Research Institute, Universidad Nacional Toribio Rodríguez de Mendoza de Amazonas (UNTRM), Amazonas, Peru, for their support and contributions throughout the completion of this research.

## Informed consent

It was not necessary to obtain informed consent in this Study.

## Author contributions

**Víctor Juan Vera-Ponce:** Methodology, Data Analysis, Writing – Review & Editing.

**Fiorella E. Zuzunaga-Montoya:** Conceptualization, Data Analysis, Writing – Review & Editing.

**Joan A. Loayza-Castro:** Data Analysis, Methodology, Validation, Writing – Original Draft.

**Luisa Erika Milagros Vásquez Romero:** Data Analysis, Methodology, Validation, Writing – Original Draft.

**Cori Raquel Iturregui Paucar:** Validation, Project Administration, Visualization, Writing – Review & Editing.

**Enrique Vigil-Ventura:** Supervision, Methodology, Funding Acquisition, Writing – Review & Editing.

## Data availability

The data supporting the findings of this study can be accessed at the following link: https://proyectos.inei.gob.pe/microdatos/

---

**Figure 1.**
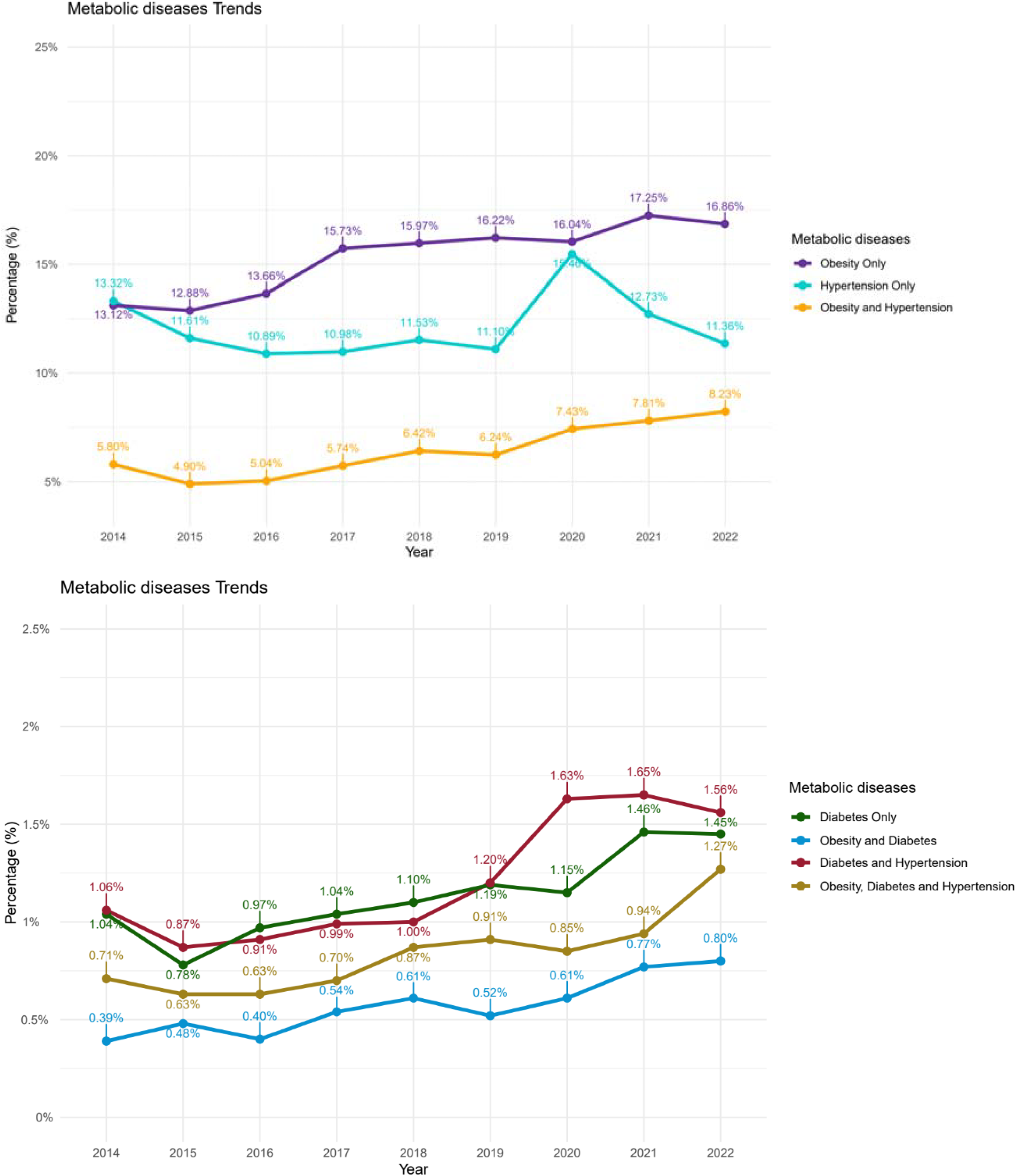
The trend from 2014 to 2022 of HTN, T2DM, Obesity, and their combinations.

## Bibliographic References

1. Non communicable diseases [Internet]. [citado el 23 de febrero de 2024]. Disponible en: https://www.who.int/news-room/fact-sheets/detail/noncommunicable-diseases

2. Piché M-E, Tchernof A, Després J-P. Obesity Phenotypes, Diabetes, and Cardiovascular Diseases. Circ Res. 2020;126(11):1477–500. doi:10.1161/CIRCRESAHA.120.316101

3. Bozkurt B, Aguilar D, Deswal A, Dunbar SB, Francis GS, Horwich T, et al. Contributory Risk and Management of Comorbidities of Hypertension, Obesity, Diabetes Mellitus, Hyperlipidemia, and Metabolic Syndrome in Chronic Heart Failure: A Scientific Statement From the American Heart Association. Circulation. 2016;134(23):e535–78. doi:10.1161/CIR.0000000000000450

4. Abarca-Gómez L, Abdeen ZA, Hamid ZA, Abu-Rmeileh NM, Acosta-Cazares B, Acuin C, et al. Worldwide trends in body-mass index, underweight, overweight, and obesity from 1975 to 2016: a pooled analysis of 2416 population-based measurement studies in 128…9 million children, adolescents, and adults. The Lancet. 2017;390(10113):2627–42. doi:10.1016/S0140-6736(17)32129-3

5. NCD Risk Factor Collaboration (NCD-RisC). Worldwide trends in hypertension prevalence and progress in treatment and control from 1990 to 2019: a pooled analysis of 1201 population-representative studies with 104 million participants. Lancet. 2021;398(10304):957–80. doi:10.1016/S0140-6736(21)01330-1

6. NCD Risk Factor Collaboration (NCD-RisC). Worldwide trends in diabetes since 1980: a pooled analysis of 751 population-based studies with 4.4 million participants. Lancet. 2016;387(10027):1513–30. doi:10.1016/S0140-6736(16)00618-8

7. Aberhe W, Mariye T, Bahrey D, Zereabruk K, Hailay A, Mebrahtom G, et al. Prevalence and factors associated with uncontrolled hypertension among adult hypertensive patients on follow-up at Northern Ethiopia, 2019: cross-sectional study. Pan Afr Med J. 2020;36:187. doi:10.11604/pamj.2020.36.187.23312

8. Apidechkul T. Prevalence and factors associated with type 2 diabetes mellitus and hypertension among the hill tribe elderly populations in northern Thailand. BMC Public Health. 2018;18(1):694. doi:10.1186/s12889-018-5607-2

9. Usui I. Common metabolic features of hypertension and type 2 diabetes. Hypertens Res. 2023;46(5):1227–33. doi:10.1038/s41440-023-01233-x

10. Walther D, Curjuric I, Dratva J, Schaffner E, Quinto C, Schmidt-Trucksäss A, et al. Hypertension, diabetes and lifestyle in the long-term - Results from a Swiss population-based cohort. Prev Med. 2017;97:56–61. doi:10.1016/j.ypmed.2016.12.016

11. Jobe M, Mactaggart I, Bell S, Kim MJ, Hydara A, Bascaran C, et al. Prevalence of hypertension, diabetes, obesity, multimorbidity, and related risk factors among adult Gambians: a cross-sectional nationwide study. Lancet Glob Health. 2024;12(1):e55–65. doi:10.1016/S2214-109X(23)00508-9

12. Price AJ, Crampin AC, Amberbir A, Kayuni-Chihana N, Musicha C, Tafatatha T, et al. Prevalence of obesity, hypertension, and diabetes, and cascade of care in sub-Saharan Africa: a cross-sectional, population-based study in rural and urban Malawi. Lancet Diabetes Endocrinol. 2018;6(3):208–22. doi:10.1016/S2213-8587(17)30432-1

13. Alcalde-Rabanal JE, Orozco-Núñez E, Espinosa-Henao OE, Arredondo-López A, Alcayde-Barranco L. The complex scenario of obesity, diabetes and hypertension in the area of influence of primary healthcare facilities in Mexico. PLoS One. 2018;13(1):e0187028. doi:10.1371/journal.pone.0187028

14. Gómez-Morales GB, Rosas-Torres BS, Hernández-Jiménez WJ, Mattenberger-Cantú E, Vargas-Villarreal J, Almanza-Reyes H, et al. Prevalence of obesity, diabetes and hypertension in immigrant populations in northeastern Mexico. Front Public Health. 2023;11:1220753. doi:10.3389/fpubh.2023.1220753

15. Bernabé-Ortiz A, Carrillo-Larco RM, Gilman RH, Checkley W, Smeeth L, Miranda JJ, et al. Contribution of modifiable risk factors for hypertension and type-2 diabetes in Peruvian resource-limited settings. J Epidemiol Community Health. 2016;70(1):49–55. doi:10.1136/jech-2015-205988

16. Carrillo-Larco RM, Bernabé-Ortiz A, Pillay TD, Gilman RH, Sanchez JF, Poterico JA, et al. Obesity risk in rural, urban and rural-to-urban migrants: prospective results of the PERU MIGRANT study. Int J Obes (Lond). 2016;40(1):181–5. doi:10.1038/ijo.2015.140

17. PERÚ Instituto Nacional de Estadística e Informática [Internet]. [citado el 30 de noviembre de 2021]. Disponible en: http://iinei.inei.gob.pe/microdatos/

18. Von Elm E G. Altman D, Egger M J. Pocock S C. Gotzsche P P. Vandenbrouckef J. Declaración de la Iniciativa STROBE (Strengthening the Reporting of Observational studies in Epidemiology): directrices para la comunicación de estudios observacionales. 22(2):144–50. doi:https://www.equator-network.org/wp-content/uploads/2015/10/STROBE_Spanish.pdf

19. NCD Risk Factor Collaboration (NCD-RisC). Worldwide trends in blood pressure from 1975 to 2015: a pooled analysis of 1479 population-based measurement studies with 19…1 million participants. Lancet. 2017;389(10064):37–55. doi:10.1016/S0140-6736(16)31919-5

20. PERU MIGRANT Study | Baseline dataset [Internet]. figshare; 2016 [citado el 14 de marzo de 2021]. doi:10.6084/m9.figshare.3125005.v1

21. Banegas JR, López-García E, Graciani A, Guallar-Castillón P, Gutierrez-Fisac JL, Alonso J, et al. Relationship between obesity, hypertension and diabetes, and health-related quality of life among the elderly. Eur J Cardiovasc Prev Rehabil. 2007;14(3):456–62. doi:10.1097/HJR.0b013e3280803f29

22. Barbaresko J, Rienks J, Nöthlings U. Lifestyle Indices and Cardiovascular Disease Risk: A Meta-analysis. American Journal of Preventive Medicine. 2018;55(4):555–64. doi:10.1016/j.amepre.2018.04.046

23. Valenzuela PL, Santos-Lozano A, Castillo-García A, Ruilope LM, Lucia A. Diabetes, Hypertension, and the Mediating Role of Lifestyle: A Cross-Sectional Analysis in a Large Cohort of Adults. Am J Prev Med. 2022;63(1):e21–9. doi:10.1016/j.amepre.2022.01.014

24. Achila OO, Araya M, Berhe AB, Haile NH, Tsige LK, Shifare BY, et al. Metabolic syndrome, associated factors and optimal waist circumference cut points: findings from a cross-sectional community-based study in the elderly population in Asmara, Eritrea. BMJ Open. 2022;12(2):e052296. doi:10.1136/bmjopen-2021-052296

25. Ansarimoghaddam A, Adineh HA, Zareban I, Iranpour S, HosseinZadeh A, Kh F. Prevalence of metabolic syndrome in Middle-East countries: Meta-analysis of cross-sectional studies. Diabetes Metab Syndr. 2018;12(2):195–201. doi:10.1016/j.dsx.2017.11.004

26. Dadwani RS, Skandari MR, GoodSmith MS, Phillips LS, Rhee MK, Laiteerapong N. Alternative type 2 diabetes screening tests may reduce the number of U.S. adults with undiagnosed diabetes. Diabet Med. 2020;37(11):1935–43. doi:10.1111/dme.14330

27. Guirguis-Blake JM, Evans CV, Webber EM, Coppola EL, Perdue LA, Weyrich MS. Screening for Hypertension in Adults: Updated Evidence Report and Systematic Review for the US Preventive Services Task Force. JAMA. 2021;325(16):1657–69. doi:10.1001/jama.2020.21669

28. Teck J. Diabetes-Associated Comorbidities. Prim Care. 2022;49(2):275–86. doi:10.1016/j.pop.2021.11.004

29. Gheorghe A, Griffiths U, Murphy A, Legido-Quigley H, Lamptey P, Perel P. The economic burden of cardiovascular disease and hypertension in low- and middle-income countries: a systematic review. BMC Public Health. 2018;18(1):975. doi:10.1186/s12889-018-5806-x

30. Oyando R, Barasa E, Ataguba JE. Socioeconomic Inequity in the Screening and Treatment of Hypertension in Kenya: Evidence From a National Survey. Front Health Serv. 2022;2:786098. doi:10.3389/frhs.2022.786098

31. Hoffman DJ, Powell TL, Barrett ES, Hardy DB. Developmental origins of metabolic diseases. Physiological Reviews. 2021;101(3):739–95. doi:10.1152/physrev.00002.2020

32. Faulkner JL, Belin de Chantemèle EJ. Sex Differences in Mechanisms of Hypertension Associated With Obesity. Hypertension. 2018;71(1):15–21. doi:10.1161/HYPERTENSIONAHA.117.09980

33. Stanhewicz AE, Wenner MM, Stachenfeld NS. Sex differences in endothelial function important to vascular health and overall cardiovascular disease risk across the lifespan. Am J Physiol Heart Circ Physiol. 2018;315(6):H1569–88. doi:10.1152/ajpheart.00396.2018

34. Hiramatsu Y, Ide H, Furui Y. Differences in the components of metabolic syndrome by age and sex: a cross-sectional and longitudinal analysis of a cohort of middle-aged and older Japanese adults. BMC Geriatr. 2023;23:438. doi:10.1186/s12877-023-04145-0

35. Kalish VB. Obesity in Older Adults. Prim Care. 2016;43(1):137–44, ix. doi:10.1016/j.pop.2015.10.002

36. Saha A, Mandal B, Muhammad T, Barman P, Ahmed W. Gender-specific determinants of overweight and obesity among older adults in India: evidence from a cross-sectional survey, 2017-18. BMC Public Health. 2023;23(1):2313. doi:10.1186/s12889-023-17156-8

37. Lamelas P, Diaz R, Orlandini A, Avezum A, Oliveira G, Mattos A, et al. Prevalence, awareness, treatment and control of hypertension in rural and urban communities in Latin American countries. Journal of Hypertension. 2019;37(9):1813. doi:10.1097/HJH.0000000000002108

38. Miranda JJ, Gilman RH, García HH, Smeeth L. The effect on cardiovascular risk factors of migration from rural to urban areas in Peru: PERU MIGRANT Study. BMC Cardiovasc Disord. 2009;9:23. doi:10.1186/1471-2261-9-23

39. Poterico JA, Stanojevic S, Ruiz-Grosso P, Bernabe-Ortiz A, Miranda JJ. The Association Between Socioeconomic Status and Obesity in Peruvian Women. Obesity. 2012;20(11):2283–9. doi:10.1038/oby.2011.288

40. Liao C, Gao W, Cao W, Lv J, Yu C, Wang S, et al. Association of Educational Level and Marital Status With Obesity: A Study of Chinese Twins. Twin Res Hum Genet. 2018;21(2):126–35. doi:10.1017/thg.2018.8

41. Laaksonen M, Talala K, Martelin T, Rahkonen O, Roos E, Helakorpi S, et al. Health behaviours as explanations for educational level differences in cardiovascular and all-cause mortality: a follow-up of 60D000 men and women over 23 years. European Journal of Public Health. 2008;18(1):38–43. doi:10.1093/eurpub/ckm051

42. AlKalbani SR, Murrin C. The association between alcohol intake and obesity in a sample of the Irish adult population, a cross-sectional study. BMC Public Health. 2023;23(1):2075. doi:10.1186/s12889-023-16946-4

43. Okojie OM, Javed F, Chiwome L, Hamid P, Okojie OM, Javed F, et al. Hypertension and Alcohol: A Mechanistic Approach. Cureus [Internet]. 2020 [citado el 22 de febrero de 2024];12(8). doi:10.7759/cureus.10086

44. Andriani H, Kosasih RI, Putri S, Kuo H-W. Effects of changes in smoking status on blood pressure among adult males and females in Indonesia: a 15-year population-based cohort study. BMJ Open. 2020;10(4):e038021. doi:10.1136/bmjopen-2020-038021

45. Taype-Rondan A, Bernabe-Ortiz A, Alvarado GF, Gilman RH, Smeeth L, Miranda JJ. Smoking and heavy drinking patterns in rural, urban and rural-to-urban migrants: the PERU MIGRANT Study. BMC Public Health. 2017;17:165. doi:10.1186/s12889-017-4080-7

46. Bouabdallaoui N, Messas N, Greenlaw N, Ferrari R, Ford I, Fox KM, et al. Impact of smoking on cardiovascular outcomes in patients with stable coronary artery disease. Eur J Prev Cardiol. 2021;28(13):1460–6. doi:10.1177/2047487320918728

47. Ellegaard PK, Poulsen HE. Tobacco smoking and oxidative stress to DNA: a meta-analysis of studies using chromatographic and immunological methods. Scand J Clin Lab Invest. 2016;76(2):151–8. doi:10.3109/00365513.2015.1127407

48. Sattar N, Preiss D. Reverse Causality in Cardiovascular Epidemiological Research: More Common Than Imagined? Circulation. 2017;135(24):2369–72. doi:10.1161/CIRCULATIONAHA.117.028307

